# Post-COVID-Syndrome patients might overestimate own cognitive impairment

**DOI:** 10.1101/2024.10.07.24314894

**Authors:** Sofia Wöhrstein, Tamara Matuz, Lilli Rötzer, Hans-Otto Karnath

**Author notes:** Correspondence should be addressed to: Prof. Dr. Dr. Hans-Otto Karnath, Center of Neurology, University of Tübingen, Hoppe-Seyler-Straße 3, D – 72072 Tübingen, Germany.

## Abstract

After a COVID-19 infection, some patients experience long-term consequences known as Post-Covid Syndrome, which often includes cognitive impairment. We investigated the congruence between subjectively experienced and objectively measured cognitive deficits after a COVID-19 infection in an unselected, successively admitted cohort of 38 patients reporting subjective cognitive complaints (SCC). We employed a comprehensive neuropsychological test battery to assess objective cognitive impairment across various cognitive domains. Three different cut-off criteria were applied commonly used in the literature to define objective neurocognitive disorder (NCD). We observed a notably low congruence between SCC and NCD in Post-Covid Syndrome, regardless of the cut-off criterion. Depending on the cognitive domain, only 5% to maximally 36% of the SCC could be objectified. One possible explanation for this discrepancy could be the high rate of depressive symptoms observed in the group of patients studied, which may negatively influence the perception of own cognitive abilities. These findings emphasize the need for careful evaluation of SCC in Post-Covid Syndrome and suggest that treating depressive symptoms may also alleviate some of the perceived cognitive deficits.

## Introduction

Some patients who have been infected with severe acute respiratory syndrome coronavirus type 2 (SARS-CoV-2) experience long-term effects following their infection [1–3]. The terminology used for this condition varies and includes different terms: e.g. Long COVID, Post-COVID symptoms, Post-COVID syndrome, Post-COVID condition, post-acute COVID [3–6]. In line with the definition of COVID-19 long term effects proposed by the National Institute for Health and Care Excellence [NICE; 7], which was also incorporated by the latest German S1 guideline for Long-/Post-COVID [5], we use the term *Post-COVID Syndrome* (PCS) in the following to address symptoms that (i) develop during or after COVID-19 infection, (ii) continue for more than 12 weeks, and (iii) cannot be explained by any other disease or diagnosis.

Around 65 million individuals worldwide are estimated to be affected by PCS, while cases are still increasing [4]. In addition to the respiratory/pulmonary sequelae, fatigue, decreased exercise tolerance, headache, anxiety, depression, insomnia, and “brain fog” were frequently reported as persistent symptoms [8, 9]. Beyond, numerous studies have reported cognitive impairments related to PCS [9–13]. Many of these studies rely on either self-reported subjective cognitive deficits in various forms or on objective results from short screening instruments such as the Montreal-Cognitive-Assessment [MoCA; 14] or the Mini-Mental-State-Examination [MMSE; 15].

A significantly smaller amount of research has focused on characterizing cognitive performance in PCS using comprehensive test batteries and clinical diagnostic criteria. They have identified deficits in attention, memory, and executive functioning following SARS-CoV-2 infection [11, 16–18]. However, the incidence rates of global cognitive impairment or of domain specific cognitive deficits fluctuate due to methodological variations regarding data collection, sample characteristics, disease severity, comorbidities, time span between infection and symptoms, and not at least due to different definitions of cognitive impairment and methods of assessment. In a meta-analysis, Ceban et al. [8] reported that 20% of the population examined in 43 studies showed cognitive impairment 12 or more weeks after COVID-19 infection. Another systematic review based on 66 studies reported incidence rates ranging from no cognitive impairment to 78% prevalence in at least one cognitive domain [19].

Given these heterogeneous rates of objective cognitive deficits persisting after COVID-19 infection and the lack of unanimous classification criteria reported to date, the present study (i) analyzes possible differences between subjective complaints and objective test results and (ii) compares the impact of different diagnostic criteria for objective cognitive disorders used in the literature. Understanding the interplay between subjectively perceived deficits in different cognitive domains and objective cognitive performance is crucial for better assessment as well as treatment options and rehabilitative measures, ultimately leading to effective patient support and a better quality of life [20–22]. To this end, we examined cognitive performance in an unselected, successively admitted group of PCS patients using comprehensive neuropsychological tests. Over a span of more than three years, these patients visited our Center for Neurology at the University Hospital Tübingen, Germany, because of subjectively perceived cognitive complaints.

## Methods

### Participants

All patients (N = 49) who were referred to the Division of Neuropsychology at the Center of Neurology at Tübingen University Hospital between 10/2020 and 01/2024 for a neuropsychological evaluation of subjectively perceived cognitive deficits (subjective cognitive complaints, SCC) after COVID-19 infection (diagnosis included PCR and/or rapid tests) were screened. In accordance with the German S1 guideline for PCS [5], they were included in the present study if their SCC (i) persisted for at least 12 weeks after their COVID-19 infection, (ii) could not be explained by other diagnosed neurological and/or current psychiatric disorders, and (iii) did not occur after COVID-19 vaccination. These inclusion criteria were met by 36 patients (21 females) aged 19 to 61 years (*M* = 47.9; *SD* = 11.6) in whom the COVID-19 infection did not require hospitalization, as well as two additional female patients aged 42 and 50 years who required hospitalization during their acute COVID-19 infection. Eleven patients were excluded (nine of them experienced a COVID-19 infection without hospitalization; two with hospitalization). For the non-hospitalized group of the final patient sample, the mean time between COVID-19 infection and neuropsychological examination was 16.1 months (*SD* = 8.3); the two hospitalized patients were examined 11 and 16 months after their respective COVID-19 infections. The study was approved by the Ethics Committee of the Medical Faculty of the University of Tübingen, Germany (authorization number 233/2024BO2).

### Neuropsychological Evaluation

The neuropsychological evaluation was conducted by a trained clinical neuropsychologist (T.M.) and lasted between 90 and 120 minutes. After a very thorough anamnesis, which served to categorize the SCC, the following neuropsychological tests were performed, all in their German version: verbal learning and memory ability test [Verbaler Lern-und Merkfähigkeitstest (VLMT); 23], Wechsler Memory Scale – Fourth Edition [WMS-IV; 24], Regensburg word fluency test [Regensburger Wortflüssigkeits-Test (RWT); 25], Tower of London [TL-D; 26], Nürnberg age inventory [Nürnberger-Alters-Inventar (NAI); 27], attention test battery [Testbatterie zur Aufmerksamkeitsprüfung (TAP); 28], and the Rey-Osterrieht Complex Figure Test [ROCFT; 29, 30]. Test performance was expressed in percentile ranks according to the respective normative test data (adjusted for age, education, and/or sex). In addition, a depression screening was performed using the long version of the general depression scale [Allgemeine Depressionsskala - lange Fassung (ADS-L); 31].

### Data Analysis

The percentile ranks from the individual subtests of the neuropsychological test battery were transformed into standardized z-scores using *psychometrica* [32]; all further analyses were performed using *R Studio* [R version 4.4.0; 33]. Following the Diagnostic and Statistical Manual of Mental Disorders [DSM-5; 34], objective cognitive impairment, i.e. a neurocognitive disorder (NCD), was defined as a z-score below one standard deviation (SD) from the respective z-score mean. If the patient had a z-score below one but less than two SDs below the mean, his/her impairment was categorized as ‘mild’. If the z-score was lower than two SDs, the impairment was categorized as ‘major’. Based on the DSM-5 criteria for NCD, we constructed five cognitive domains from the subtests of the aforementioned neuropsychological test battery: 1) Attention, 2) Memory and Learning, 3) Executive Function, 4) Word Fluency, and 5) Visual Reproduction. Table 1 shows which subtests were assigned to which domains. In parallel, we also categorized patient-reported symptoms and deficits (SCC) in the same five cognitive domains.

**Table 1.** Assignment of (sub-)tests of the neuropsychological evaluation to cognitive domains.

| Domain | Cognitive task | Neuropsychological (sub-)test |
| --- | --- | --- |
| **Attention** | Divided attention | TAP divided total |
|  | Tonic attention | TAP tonic alertness |
|  | Phasic attention | TAP phasic K value |
|  | Flexible attention | TAP flexible |
| **Memory and Learning** | Verbal learning | VLMT sum rounds 1-5 /<br>WMS-IV logical memory |
|  | Verbal recall immediate | VLMT difference round 5-round 6 /<br>WMS-IV logical memory |
|  | Verbal recall delayed | VLMT difference round 5-round 7 /<br>WMS-IV logical memory |
|  | Verbal recognition | VLMT recognition /<br>WMS-IV logical memory |
|  | Short term memory | WMS-IV digit span forward |
| **Executive Function** | Working memory | NAI /<br>WMS-IV digit span backward |
|  | Categorical switch word fluency | RWT sports/fruits |
|  | Categorical switch word fluency | RWT G/R-words |
|  | Cognitive planning | TL-D |
| **Word Fluency** | Semantic word fluency | RWT animals |
|  | Phonematic word fluency | RWT P-words |
| **Visual Reproduction** | Visual reproduction immediate | ROCFT /<br>WMS-IV figure |
|  | Visual reproduction delayed | ROCFT /<br>WMS-IV figure |
|  | Visual recognition | ROCFT /<br>WMS-IV figure |

*Abbreviations:* NAI = Nürnberg age inventory [27]; ROCFT = Rey-Osterrieth Complex Figure Test [29, 30]; RWT = Regensburg word fluency test [25]; TAP = attention test battery [28]; TL-D = Tower of London [26]; VLMT = verbal learning and memory ability test [23]; WMS-IV = Wechsler Memory Scale – Fourth Edition [24].

To date, various analysis criteria as well as NCD cut-off scores have been used in the literature to analyze and classify cognitive domains. To compare the effects of using these different approaches, we analyzed our data according to the following three criteria. The first and most liberal criterion “*subtests overall*” [e.g., 11] refers to the standardized z-scores for each subtest. In this case, if a patient had an impairment in at least two subtests of the total of 18 subtests *across all five cognitive domains* (see above), he/she was classified as having a NCD. A second criterion “*mean domain*” [e.g., 17] is based on the mean z-score per cognitive domain, i.e. the averaged z-score across all subtests that make up a particular cognitive domain. In the third and most conservative criterion “*subtests domain*” [cf. 34], we considered the individual z-scores for each subtest *per cognitive domain*. If at least two subtests in a domain were between one and two SD below the mean of the subtest, the domain was classified as ‘mildly’ impaired; if two subtests were more than two SD below the mean of the subtest, the domain was classified as ‘majorly’ impaired. If only one subtest was at least one SD below the mean, this domain was classified as ‘not impaired’. According to the DSM-5 guidelines, only one of the five cognitive domains had to be impaired to be classified as a mild or major general NCD.

## Results

### COVID-19 infections without hospitalization

#### Subjective cognitive complaints (SCC)

Most patients from the group without hospitalization reported SCC in the domain of *Attention*, followed by *Memory and Learning*, *Executive Function*, and *Word Fluency*, while none reported symptoms or deficits in *Visual Reproduction* (Fig. 1). In addition to complaints in these five cognitive domains, 41.7% of the patient sample reported headaches and 91.7% “rapid exhaustion” and/or “tiredness” in everyday life.

**Fig. 1.**
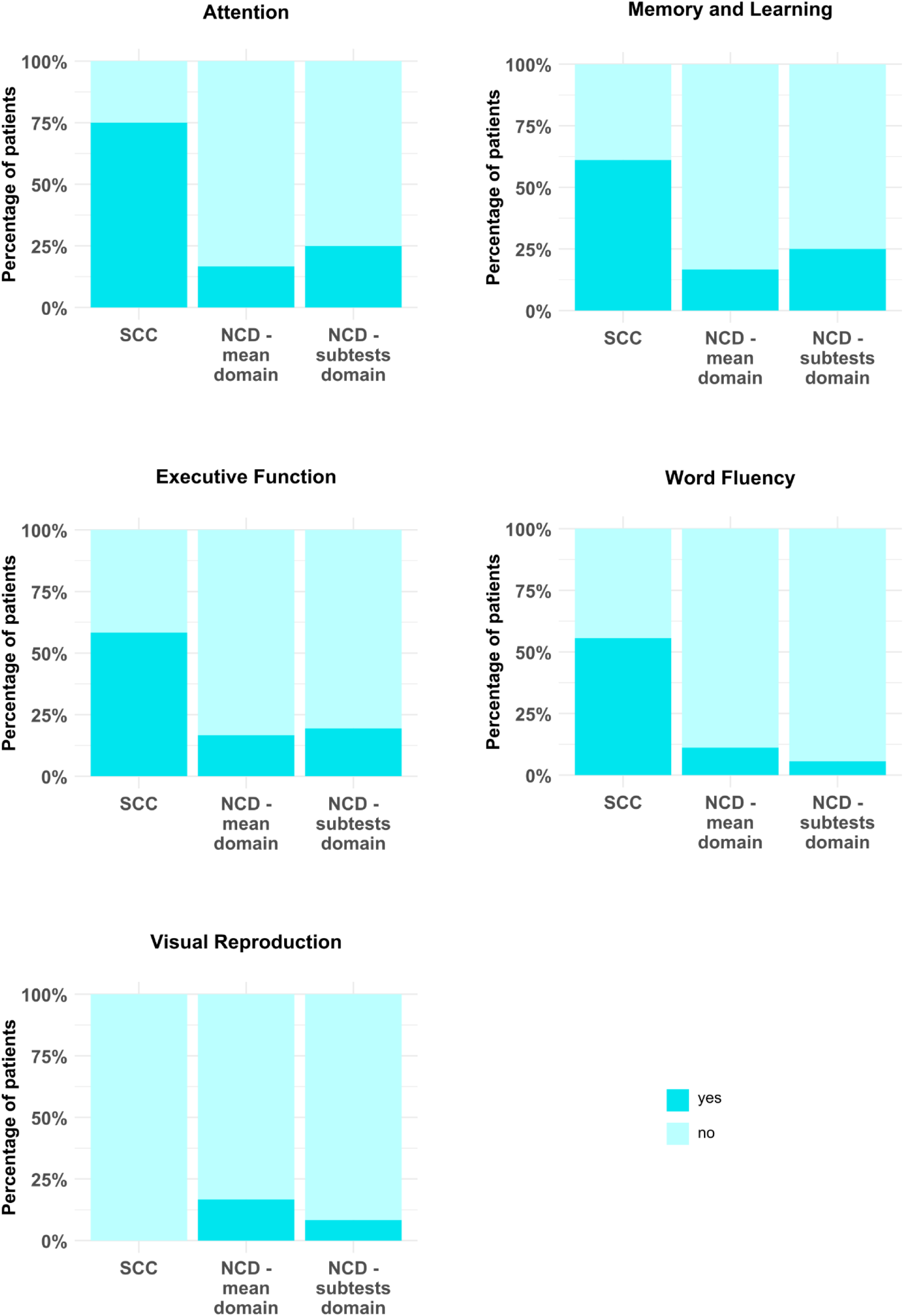
Subjective cognitive complaints (SCC) and objectively measured neurocognitive disorders (NCD) in the patient group without hospitalization (N=36). The barplots show the percentages of patients who reported SCC in the respective cognitive domain as well as the percentages of patients who showed objective NCD with respect to the criteria *‘mean domain’* and *‘subtests domain’*. Mild and major NCD have been combined. (Note that the criterion “*subtests overall*” could not be illustrated in this figure since it did not take into account each domain separately.) No patient reported deficits in the domain *Visual Reproduction*.

#### Objective neurocognitive disorder (NCD)

Two criteria for objective NCD frequencies (*‘mean domain’* and *‘subtests domain’)* allowed to take into account each domain separately and thus could be illustrated in comparison with the patients’ SCC (cf. Fig. 1). Numerically, in the domains *Attention*, *Memory and Learning*, as well as *Executive Function*, slightly fewer patients with an NCD were identified using the *‘mean domain’* criterion compared to *‘subtests domain’* criterion. However, in the domains of *Word Fluency* and *Visual Reproduction*, the *‘subtests domain’* criterion identified a slightly higher number of affected patients. Detailed neuropsychological (sub-)test results for each domain according to all three criteria are provided in Table S1 in the supplement.

To compare the effects of the different NCD cut-off criteria on the NCD prevalence rate, we analyzed the same data set with respect to three different criteria. With the first and most liberal criterion (two subtest over all domains [*‘subtests overall’*]) 21 patients (58.3%) were classified having mild and two patients (5.6%) having major NCD, while 13 (36.1%) had no NCD. Using the second criterion (mean z-scores per domain [*‘mean domain’*]), 13 patients (36.1%) were classified with mild and two patients (5.6%) with major NCD, while 21 patients (58.3%) had no NCD. Finally, using the third criterion by looking at two subtests per domain [*‘subtests domain’*], 13 patients (36.1%) reached the cut-off for mild and two patients (5.5%) major NCD, while 21 (58.3%) had no NCD. Statistical comparison of the three different NCD cut-off criteria revealed that there was no significant difference in terms of the proportion of patients with NCD (*χ^2^*[2, N=36] = 4.74, *p* = .093; *φ* = 0.36).

Figure 2 illustrates all demographical variables for the group of non-hospitalized patients with and without objective NCD for each combination of the three NCD cut-off criteria. A table containing all detailed demographical information can be found in the supplementary material (Table S2). The duration of education for this group, measured in school years plus years of study or vocational training, was 14.3 years (*SD* = 2.4, range 9 – 19). The patient’s depression scores were above average, quantified in percentile ranks with a median of 86 (*IQR* = 24.5).

**Fig. 2.**
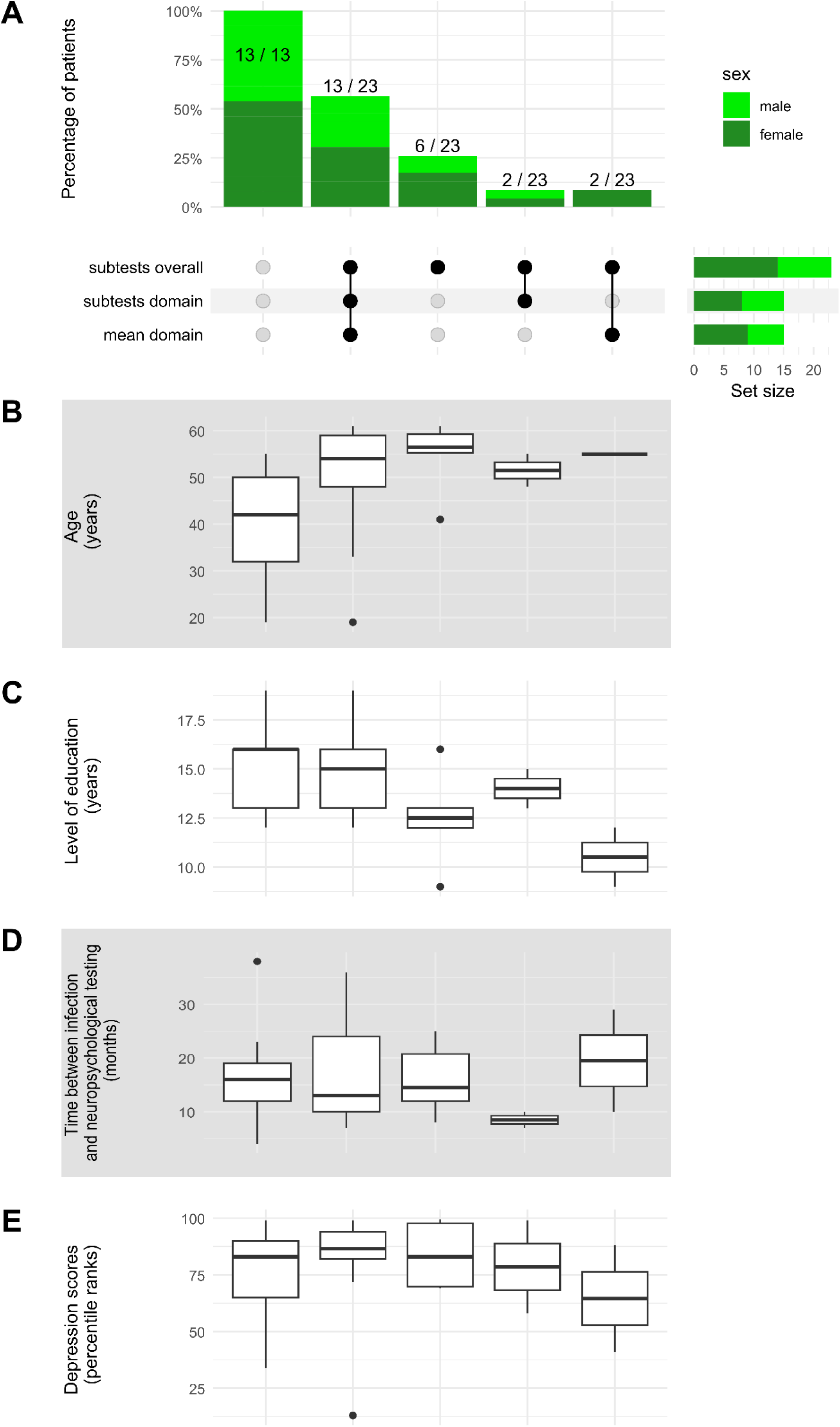
Comparison of the three neurocognitive disorder (NCD) cut-off criteria *‘subtests overall’*, *‘subtests domain’* and *‘mean domain’* using a complex UpSet plot. Mild and major NCD have been combined. **A) Sex**: The UpSet plot shows the percentage of patients for each combination of NCD cut-off criteria, broken down by sex. The lower right corner shows the number of patients per criterion who were classified as having a NCD. **B) Age**: Boxplots with the median age of the respective patient group for each combination of NCD cut-off criteria. **C) Level of education**: Boxplots with the median level of education of the respective patient group for each combination of NCD cut-off criteria. The level of education has been measured in school years plus years of study or vocational training. **D) Time between COVID-19 infection and neuropsychological testing**: Boxplots with the median time interval in months between the COVID-19 infection and the neuropsychological evaluation of the respective patient group for each combination of NCD cut-off criteria. **E) Depression**: Boxplots with the median depression scores, illustrated in percentile ranks, of the respective patient group for each combination of NCD cut-off criteria.

For the group of patients with NCD (in either of the three criteria), sex did not differ significantly between ‘*subtests overall*’, ‘*mean domain*’ and ‘*subtests domain*’ (*χ^2^* [2, N=31] = 0.23, *p* = .891; *η^2^* = 0.08). Also age did not differ significantly between the three criteria (*χ^2^* [2] = 0.30, *p* = .859; *η^2^* = -0.03), nor did the time between COVID-19 infection and neuropsychological examination (*χ^2^* [2] = 0.29, *p* = .866; *η^2^* = -0.03), the level of education (*χ^2^* [2] = 1.35, *p* = .510; *η^2^* = -0.01), or the depression scores (*χ^2^* [2] = 0.04, *p* = .982; *η^2^* = -0.04). For the group of patients without NCD, sex also did not differ significantly between ‘*subtests overall*’, ‘*mean domain*’ and ‘*subtests domain*’ (*χ^2^* [2, N=32] = 0.23, *p* = .892; *η^2^* = 0.08), also age did not differ between the three criteria (*χ^2^* [2] = 2.77, *p* = .0.250; *η^2^* = 0.02), as well as the time between COVID-19 infection and neuropsychological examination (*χ^2^* [2] = 0.21, *p* = .901; *η^2^* = -0.03), the level of education (*χ^2^* [2] = 1.69, *p* = .429; *η^2^* = -0.01) or depression scores (*χ^2^* [2] = 0.28, *p* = .871; *η^2^* = -0.03).

#### Comparing SCC and NCD

To investigate how well the objectively assessed cognitive deficits matched the subjectively reported cognitive complaints, we determined whether or not the subjectively reported deficit of a patient fitted with the objectively measured deficit (or non-existent deficit) of the same patient. To this end, we measured the congruence on a global level, meaning that we looked at the congruence of a patient for each cognitive domain and then averaged it. As a result, we obtained global congruence percentages (50% congruency indicates that subjects had the same number of matches as non-matches between SCC and NCD). Overall, the comparison between subjective complaints and objective findings did not show a high congruence (Fig. 3). More than half of our patient sample was not congruent regarding SCC and NCD, i.e., they had less than 50% congruency, and only one patient from our sample showed global congruence higher than 50% (Fig. 3). In the non-congruent or low-congruent part of our patient sample (0% or 25% congruence), 59.4% were female. The mean age of all (male and female) patients in this subgroup was 47 years (*SD* = 12), the level of education 14.1 years (*SD* = 2.5) and the time between their COVID-19 infection and the neuropsychological examination 16.3 months (*SD* = 8.4). Interestingly, the depression scores of all (male and female) patients in this subgroup were in the above-average range (i.e., ≥ percentile rank of 84) in 88.9% of cases; the median depression score was a percentile rank of 87 (*IQR* = 27).

**Fig. 3.**
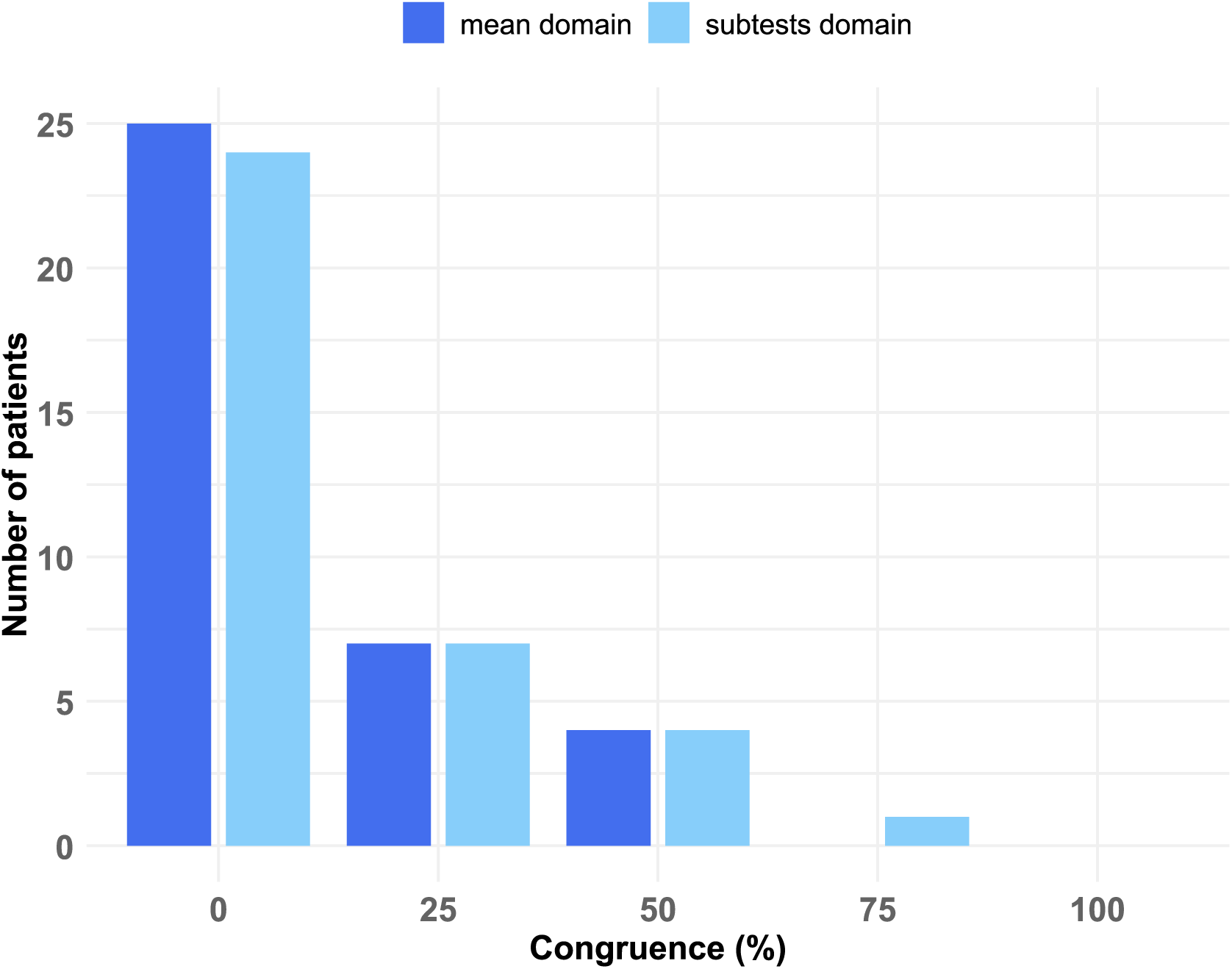
Global congruence between subjective cognitive complaints (SCC) and objective neurocognitive disorders (NCD) over four cognitive domains (without *Visual Reproduction* since no patient reported SCC in this domain). The barplot illustrates the number of patients who obtained different congruence percentages (measured as the average of congruence between four cognitive domains), separately for our NCD cut-off criteria *mean domain* and *subtests domain*.

To go into more detail, we also determined the congruence between SCC and NCD separately for each cognitive domain (Tab. 3). In the majority of cases (between 64% and 95%, depending on the respective domain and criterion), the subjectively reported deficit in a specific cognitive domain could not be verified by the neuropsychological test results of this domain. The congruence between SCC and objective NCD was the highest in the domains *Memory and Learning* as well as *Attention* (cf. Tab. 3), while it was lowest in the cognitive domains *Executive Function* and *Word Fluency*.

**Table 3.** Domain-specific congruence (in percent) of objective cognitive impairment (NCD) found in the non-hospitalized patients (N=36) who subjectively reported cognitive impairment (SCC) in that domain.

| Cognitive domain | Congruence between SCC and NCD |  |
| --- | --- | --- |
|  | Criterion ‘Mean domain’ | Criterion ‘Subtests domain’ |
| Attention<br>N=27 reporting SCC | 18.52 % | 22.22 % |
| Memory and Learning<br>N=22 reporting SCC | 27.27 % | 36.36 % |
| Executive Function<br>N=21 reporting SCC | 9.52 % | 14.29 % |
| Word Fluency<br>N=20 reporting SCC | 10.00 % | 5.00 % |
| Visual Reproduction<br>N=0 reporting SCC | n.a. | n.a. |

*Note.* In contrast to figure 1, this table shows the domain-specific congruence between subjective cognitive complaints (SCC) and neurocognitive disorder (NCD) for each individual; i.e., we determined whether or not the subjectively reported deficit of a patient in one domain fitted with the objectively measured deficit (or non-existent deficit) of the same patient in the same domain. Values are presented for only two of the three cut-off criteria, since criterion ‘subtests overall’ is based on the number of impaired subtests from all subtests, regardless of domain. For the domain *Visual Reproduction* no congruence values can be provided because none of our patients reported symptoms or deficits in this domain. *Abbreviations:* n.a. = not applicable; NCD = objective neurocognitive disorder; SCC = subjective cognitive complaints.

On the other hand, if patients did not report SCC for a specific domain, this was consistent with our objective results in the majority of cases (67% or more, depending on the respective domain). However, some cases were also observed where the reverse was true. Patients who did not report deficits in the *Attention* domain but had objective deficits in 33.3% (*‘subtests domain’*) or 11.1% (*‘mean domain’*). With regard to the domains *Memory and Learning* 7.1% (*‘subtests domain’*) or 0% (*‘mean domain’*), *Executive Function* 26.7% (*‘subtests domain’* as well as *‘mean domain’*), and *Word Fluency* 6.3% (*‘subtests domain’*) or 12.5% (*‘mean domain’*) of patients who did not complain deficits had objective deficits in the corresponding domain.

### COVID-19 infections that required hospitalization

Only two patients in our unselected sample that fulfilled the inclusion criteria (see above) had to be hospitalized during their acute COVID-19 infection; one of them including a stay at the intensive care unit. Both patients reported SCC in only one cognitive domain each (patient 1: *Memory and Learning*, patient 2: *Word Fluency*). Furthermore, they reported ‘rapid exhaustion’ and ‘tiredness’ in everyday life but no headaches. Neither of the two hospitalized patients had a NCD for any of the three criteria (‘subtests overall’, ‘mean domain’, ‘subtests domain’). Thus, there was no domain-specific congruence of objectively measured (NCD) and subjectively reported (SCC) cognitive impairment in the two hospitalized patients. The duration of the education of the two patients was 19 and 13 years; the depression percentile ranks 86 and 34 and thus above average in one case. Detailed neuropsychological (sub-)test results are shown in Table S3 in the supplement.

## Discussion

We investigated the congruence of subjective and objective cognitive deficits after a COVID-19 infection in an unselected cohort of 38 patients with subjectively perceived cognitive complaints, successively admitted in our Center for Neurology over a period of a good three years. The cognitive domains that were subjectively most often complained about as impaired were attention, memory and executive functions, while language-related and visuo-spatial abilities were the least complained about as impaired. We compared these subjective complaints with the objective, psychometric measurement of these abilities. In order to avoid any disagreement about how cognitive impairment should be assigned when objectifying these subjectively perceived cognitive impairments, we applied three different criteria, commonly used in the literature. As expected, the more liberal criterion (“*subtests overall*”) for defining NCD resulted in the highest rate of objective NCD, whereas the two more conservative criteria (“*mean domain*” and “*subtests domain*”) resulted in lower NCD rates. However, statistical comparison showed that the NCD rates in our sample did not differ significantly. Further, our results suggest that − independent of the classification criteria − SCC were only seldom in congruence with the objectively measurable NCD. In general, SCC rates were much higher compared to the objective NCD in all investigated cognitive domains. Only 14% of our non-hospitalized patients had a congruence rate of ≥50% between their SCC report and the actually measured NCD; 86% showed either no congruence at all or one of only 25%.

Discrepancies between subjective and objective rates of cognitive impairment in PCS have also been found by other researchers. Klinkhammer et al. [35] observed that subjective “cognitive complaints […] exceed cognitive dysfunction by far”. Unfortunately, they did not provide exact percentages for the congruence between subjective complaints and objective deficits, as this comparison was not the focus of their study. Another study that investigated the congruence between self-reported cognitive impairment and objective test results reported a small correlation (determined by linear regression analysis) between subjective and objective measures [36]. However, this study relied on only three neuropsychological tests rather than an entire test battery and, more important, did not aim to identify the proportion of patients who actually have objective cognitive deficits from those who reported subjective cognitive deficits. Comparable with the present study, Schild et al. [12] used comprehensive and domain-specific neuropsychological tests and reported an overall correspondence between SCC and NCD of ∼40%, when combining their baseline and follow-up examinations. Our findings showed a comparable congruence of maximally 36% (depending on the cognitive domain) between SCC and NCD.

In addition to patients that subjectively complained about cognitive disorders that could not be objectified, we also observed patients who showed the opposite dissociation. These patients did not report SCC in a specific domain but showed objective results suggestive of a domain-related NCD. The rates of patients with objective deficits but no subjective cognitive complaints ranged from 0% to 33%, depending on the cognitive domain. The question arises as to whether these patients simply forgot to mention deficits in these domains, or whether they genuinely believe that they have no impairments in these domains. Since the procedure in our study did not include explicit asking about specific subjectively experienced deficits in each cognitive domain, it is difficult to determine the exact nature of this discrepancy. Our cohort of patients with subjectively perceived cognitive complaints was on average 48 years old and the level of education was on average 14 years. Thus, our sample was very similar to the PCS patients with subjective and/or objective cognitive deficits investigated in previous studies [12, 16, 18]. In addition, our sample consisted almost exclusively of patients with COVID-19 infections that did not require hospitalization in the acute phase, as in Fleischer et al. [38]. Remarkably, we found no significant differences in the proportion of men and women in our sample, although previous studies have suggested a higher proportion of women in PCS patients [e.g. 18, 38-42]. The high level of education in our sample, with more than half having an academic degree or even a doctorate is interesting and could possibly be explained by the fact that patients with a higher level of education may be more responsive to small cognitive changes or show a greater interest in health issues in general [43] and are more likely to present to hospitals [44]. In addition, the usual occupations in which highly educated patients work may require and consequently train (more) finely tuned cognitive resources [38]. These factors may explain why these patients are more attuned to their cognitive abilities and quicker to notice changes and subjective declines.

There is another important aspect that could underlie the present finding that the subjectively perceived cognitive deficits could not be measured objectively in the greater part of our unselected patient sample. Very clearly, the depression scores of all (male and female) patients in the group who reported SCC but did not show an objective NCD were in the above-average range in 89% of cases. This high level of depressive symptoms in our sample corresponds with previous observations of depressive symptoms or even clinical depression in PCS [41, 45–47]. However, we would like to emphasize that our study does not allow us to draw conclusions about the causality of the depressive symptoms. On the one hand, it is possible that depressive symptoms or depression cause the subjective perception of a cognitive dysfunction and/or impairment [48–51], particularly in the domains of executive function, memory and attention, which were also most affected in our patient sample. Reduced cognitive abilities are even part of the DSM-V diagnostic criteria [34] for major depressive disorder. Consequently, depressive symptoms can impair both one’s own cognitive abilities as well as their perception [52], regardless of the COVID-19 infection. Schwert et al. [52] even showed that depressed patients underestimate their cognitive abilities, which, albeit in relation to a different disease, is congruent with the results of our present study. The same was also found in another study in which subjective and objective cognitive impairment following traumatic brain injury was investigated [53]. They also observed high levels of depression in their patient sample and interpreted their results primarily by an underestimation of cognitive performance. On the other hand, however, the observed depressive symptoms could be the consequence of the COVID-19 infection itself, as depression and psychiatric diagnoses in general have an increased incidence after COVID-19 [45, 54]. The causality between the experienced cognitive impairment in PCS and the presence of depressive symptoms therefore remains plausible in both directions.

### Conclusion

In conclusion, this study showed that subjective cognitive complaints following COVID-19 infection often do not align with results from standardized neuropsychological tests. In our unselected sample, most of the patients showed either no congruence at all or one of only 25% between SCC and objectively measured NCD. The high rate of depressive symptoms in our patient sample was remarkable and might be one factor underlying this discrepancy. Addressing depressive symptoms in treatments and psychotherapy in PCS may alleviate the overall burden of suffering, ultimately leading to an improved quality of life for the affected patients.

## Data Availability

All data produced in the present study are available upon reasonable request to the authors.

## Abbreviations

COVID-19: coronavirus disease 2019
DSM-5: Diagnostic and Statistical Manual of Mental Disorders, fifth edition
IQR: interquartile range
MMSE: Mini-Mental State Examination
MoCA: Montreal Cognitive Assessment
NCD: neurocognitive disorder
PCS: Post-COVID syndrome
PR: percentile rank
SARS-CoV-2: severe acute respiratory syndrome coronavirus type 2
SCC: subjective cognitive complaints
SD: standard deviation.

## Statements and Declarations

## Acknowledgements

We would like to thank all patients who participated in our study.

## Author contributions

SW, TM and H-OK contributed to the study conception and design. TM conducted neuropsychological testing and data was prepared by LR. SW carried out the data analysis and created the figures. The first draft of the manuscript was written by SW and TM and all authors commented on previous versions of the manuscript. All authors read and approved the final manuscript.

## Ethics approval

This study was approved by the Ethics Committee of the Medical Faculty of the University of Tübingen, Germany (233/2024BO2) and performed in line with the principles of the Declaration of Helsinki (1964).

