## Supplementary Material for "Post-COVID-Syndrome patients might overestimate own cognitive impairment"

**Supplementary Tables**

**Table S1.** **Mean z-scores and standard deviations of all subtests of the neuropsychological evaluation for the group of non-hospitalized patients (N = 36).**

| **Domain** | **Mean z-score (SD)** | **Cognitive task** | **Mean z-score (SD)** | **Neuropsychological (sub)test** |
| --- | --- | --- | --- | --- |
| **Attention** | -0.31 (0.57) | Divided attention | -0.44 (0.99) | TAP divided total |
|  |  | Tonic attention | -0.85 (1.16) | TAP tonic alertness |
|  |  | Phasic attention | 0.05 (1.12) | TAP phasic K value |
|  |  | Flexible attention ^N=32^ | 0.08 (0.87) | TAP flexible |
| **Memory and Learning** | -0.30 (0.60) | Verbal learning ^N=26^ | 0.10 (0.98) | VLMT sum rounds 1-5 /  WMS-IV logical memory |
|  |  | Verbal recall immediate | -0.49 (0.89) | VLMT difference round 5-round 6 /  WMS-IV logical memory |
|  |  | Verbal recall delayed | -0.64 (0.93) | VLMT difference round 5-round 7 /  WMS-IV logical memory |
|  |  | Verbal recognition | -0.25 (0.74) | VLMT recognition / WMS-IV logical memory |
|  |  | Short term memory | 0.02 (0.99) | WMS-IV digit span forward |
| **Executive Function** | 0.05 (0.54) | Working memory ^N=35^ | -0.05 (0.88) | NAI / WMS-IV digit span backward |
|  |  | Categorical switch word fluency | 0.20 (0.87) | RWT sports/fruits |
|  |  | Categorical switch word fluency | -0.33 (0.77) | RWT G/R-words |
|  |  | Cognitive planning ^N=29^ | 0.45 (1.06) | TL-D |
| **Word Fluency** | -0.05 (0.65) | Semantic word fluency | 0.24 (0.85) | RWT animals |
|  |  | Phonematic word fluency | -0.35 (0.80) | RWT P-words |
| **Visual Reproduction ^N=35^** | 0.49 (1.21) | Visual reproduction immediate ^N=35^ | 0.63 (1.30) | ROCFT / WMS-IV figure |
|  |  | Visual reproduction delayed ^N=35^ | 0.16 (1.52) | ROCFT / WMS-IV figure |
|  |  | Visual recognition ^N=17^ | -0.02 (0.57) | ROCFT / WMS-IV figure |

*Note*. Abbreviations: n.a. = not applicable; NAI = Nürnberg age inventory [1]; NCD = neurocognitive disorder; ROCFT = Rey-Osterrieth Complex Figure Test [2, 3]; RWT = Regensburg word fluency test [4]; SD = standard deviation; TAP = attention test battery [5]; TL-D = Tower of London [6]; VLMT = verbal learning and memory ability test [7]; WMS-IV = Wechsler Memory Scale – Fourth Edition [8].

**Table S2.** **Demographical variables for the non-hospitalized patients with versus without objective neurocognitive disorder (NCD versus no NCD).**

|  | **Criterion ‘*subtests overall*’** | | | **Criterion ‘*mean domain*’** | | | **Criterion ‘*subtests domain*’** | | |
| --- | --- | --- | --- | --- | --- | --- | --- | --- | --- |
|  | **No NCD** | **Minor NCD** | **Major NCD** | **No NCD** | **Minor NCD** | **Major NCD** | **No NCD** | **Minor NCD** | **Major NCD** |
| N | 13 | 21 | 2 | 21 | 13 | 2 | 21 | 14 | 1 |
| Age, *years* (SD) | 40.92 (10.96) | 51.62 (10.71) | 54.00 (5.66) | 45.95 (11.42) | 52.00 (11.65) | 41.5 (12.02) | 46.29 (11.59) | 50.14 (12.25) | 50.00 (n.a.) |
| Sex (f/m) | 7/6 | 13/8 | 1/1 | 12/9 | 8/5 | 1/1 | 13/8 | 7/7 | 1/0 |
| Months between infection and neuropsychological exam (SD) | 16.38 (8.74) | 15.05 (8.05) | 25.00 (1.41) | 15.52 (7.89) | 15.62 (9.05) | 25.00 (1.41) | 16.57 (8.16) | 14.64 (8.48) | 26.00 (n.a.) |
| Level of education, *years* (SD) | 14.92 (2.02) | 13.52 (2.36) | 17.50 (2.12) | 14.14 (2.24) | 14.08 (2.56) | 16.50 (3.54) | 13.81 (2.52) | 14.57 (1.91) | 19.00 (n.a.) |
| Depression scores, *PR median* (IQR) | 83  (25) | 87  (25) | 82 (0) | 83 (26) | 86 (6) | 93 (0) | 83 (23) | 86.50 (14) | n.a. |

*Note*. Abbreviations: SD = standard deviation; n.a. = not applicable; f = female; m = male; PR = percentile rank; IQR = interquartile range.

**Table S3.** **Standardized z-scores of all subtests of the neuropsychological evaluation for the two hospitalized patients.**

| **Domain** | **Patient 1** | **Patient 2** | **Cognitive task** | **Patient 1** | **Patient 2** | **Neuropsychological (sub)test** |
| --- | --- | --- | --- | --- | --- | --- |
| **Attention** | 0.28 | -0.70 | Divided attention | 0.00 | -1.28 | TAP divided total |
|  |  |  | Tonic attention | 0.71 | -1.18 | TAP tonic alertness |
|  |  |  | Phasic attention | 0.31 | -0.81 | TAP phasic K value |
|  |  |  | Flexible attention | 0.10 | 0.50 | TAP flexible |
| **Memory and Learning** | -1.00 | 0.74 | Verbal learning | -1.64 | 1.64 | VLMT sum rounds 1-5 / WMS-IV logical memory |
|  |  |  | Verbal recall immediate | -0.39 | 0.67 | VLMT difference round 5-round 6 /  WMS-IV logical memory |
|  |  |  | Verbal recall delayed | -0.39 | 0.52 | VLMT difference round 5-round 7 /  WMS-IV logical memory |
|  |  |  | Verbal recognition | -1.64 | 0.84 | VLMT recognition / WMS-IV logical memory |
|  |  |  | Short term memory | -0.92 | 0.00 | WMS-IV digit span forward |
| **Executive Function** | -0.82 | 0.35 | Working memory | -1.18 | 0.67 | NAI / WMS-IV digit span backward |
|  |  |  | Categorical switch word fluency | -0.10 | 0.61 | RWT sports/fruits |
|  |  |  | Categorical switch word fluency | -1.18 | 0.77 | RWT G/R-words |
|  |  |  | Cognitive planning ^N=1^ | n.a. | -0.67 | TL-D |
| **Word Fluency** | -0.70 | 0.50 | Semantic word fluency | -0.23 | 0.74 | RWT animals |
|  |  |  | Phonematic word fluency | -1.18 | 0.25 | RWT P-words |
| **Visual Reproduction** | 0.84 | 0.41 | Visual reproduction immediate | 0.50 | 0.41 | ROCFT / WMS-IV figure |
|  |  |  | Visual reproduction delayed | 1.18 | 0.41 | ROCFT / WMS-IV figure |
|  |  |  | Visual recognition ^a^ | n.a. | n.a. | ROCFT / WMS-IV figure |

*Note*. Abbreviations: n.a. = not applicable; NAI = Nürnberg age inventory [1]; NCD = neurocognitive disorder; ROCFT = Rey-Osterrieth Complex Figure Test [2, 3]; RWT = Regensburg word fluency test [4]; SD = standard deviation; TAP = attention test battery [5]; TL-D = Tower of London [6]; VLMT = verbal learning and memory ability test [7]; WMS-IV = Wechsler Memory Scale – Fourth Edition [8]. ^a^This subtest was not performed on either of the two patients.
